# Low retention on PrEP and barriers to PrEP use among key populations in Kinshasa, DRC: a mixed method study

**DOI:** 10.1101/2024.12.18.24318849

**Authors:** Natalia Zotova, Alisho Shongo, Patricia Lelo, Nana Mbonze, Didine Kaba, Paul Ntangu, Qiuhu Shi, Adebola Adedimeji, Kathryn Anastos, Marcel Yotebieng, Viraj Patel, Jonathan Ross

## Abstract

Female sex workers (FSW) and men who have sex with men (MSM) are disproportionately affected by HIV. Oral pre-exposure prophylaxis (PrEP) has become increasingly available in African countries including the Democratic Republic of Congo (DRC). However, limited data exist on PrEP uptake and retention or on factors that affect PrEP use among FSW and MSM. This mixed-method study, conducted at KP-friendly centers in Kinshasa, DRC, aimed to identify patterns of PrEP retention and to understand underlying factors of PrEP engagement. Collected data included programmatic data, extraction of routine clinical records, and qualitative interviews with FSW and MSM. Logistic regression was used to identify factors associated with PrEP retention. Qualitative data were analyzed thematically. Findings were then synthesized. Low rates of PrEP initiation and retention were of concern. Only 25% of eligible FSW and MSM initiated PrEP in 2019-2022. Among FSW, 79% returned to the clinic for PrEP refills at 1 month, with only 15% returning for a 3-month visit. Similarly, 74% of MSM were retained at 1 month, with 10% retained at 3 months. Previous experience using PrEP was significantly associated with retention at 3 months. Qualitative analyses identified stigma, side effects, dislike of daily medication regimen, and a shortage of KP-friendly facilities as major barriers to PrEP engagement. This warrants the need for interventions to strengthen messaging about PrEP and side effects wading over time. Raising awareness among the Congolese general population may help to avoid stigmatization of PrEP users and improve PrEP acceptance among key populations at risk.

## INTRODUCTION

HIV, which remains a major public health problem, has disproportionately impacted Sub-Saharan Africa (SSA) countries. Of the estimated 39 million people living with HIV (PLWH), 25.6 million live in the SSA region. Globally, 1.3 million people contracted HIV in 2022 (1) and more than half of all new infections occurred among key populations (KPs) – female sex workers (FSW), men who have sex with men (MSM), transgender persons (TG), injection drug users, and HIV-negative partners in serodifferent partnerships. In SSA, 1 out of 4 of new infections was among KPs, their sexual partners and clients (1).

Pre-Exposure prophylaxis (PrEP) is highly effective in reducing the risk of HIV acquisition (2,3). Oral PrEP has become increasingly available in low– and middle-income countries including SSA. It was first introduced in South Africa in 2016 with initial provision at demonstration sites followed by a scaled-up provision for key populations (4). National programs to deliver PrEP to population groups at high risk of HIV including FSW and MSM are rapidly scaling up in other African countries including the Democratic Republic of Congo (DRC). Early demonstration studies among KPs in Benin (5), Kenya and Uganda (6), Senegal (7), and South Africa (8) have shown PrEP acceptability and effectiveness. However, PrEP uptake and retention among FSW and MSM in real-world settings remain suboptimal (9–16).

The HIV epidemic in Western and Central Africa differs from Eastern and Southern Africa. While HIV prevalence among the general population in Western and Central Africa continues to fall, hovering around 1% or lower, KPs are disproportionately affected by HIV (17,18). Understanding HIV prevention use and needs among KPs is needed to inform ongoing efforts to address the epidemic. Yet, few data on the PrEP continuum of care have been reported from this region. Evidence from Cameroon pointed out low rates of PrEP initiation and continuation among FSW and MSM: 45% of eligible individuals had initiated PrEP, and 37% and 19% of clients who initiated on PrEP, continued using it at 3 and 12 months, respectively (18). A study from Rwanda found similar uptake, but higher continuation rates: PrEP uptake was 46% among FSWs and 35% among MSM. Among PrEP initiators, 79% of FSW and 88% of MSM continued using it at 12 months (17).

DRC, the largest country in Central Africa, has been scaling up PrEP since 2019 focusing on key population groups at risk of HIV (19,20). While an early pilot study in 2018 (19) reported high acceptability of PrEP, no subsequent research has examined outcomes among FSW and MSM despite an estimated 60,000 individuals initiated on PrEP during this time (21). Similarly, information is scant on barriers and facilitators of PrEP use and experiences of PrEP users in this region. We conducted a mixed-method study of Congolese FSW and MSM who were eligible for PrEP to identify patterns of retention in care and understand underlying factors of PrEP engagement in a real-world setting.

## METHODS

### Study design, population, and setting

This mixed-method study used a convergent parallel design to collect, analyze, and integrate programmatic data from participating clinics and primary quantitative and qualitative data collected from FSW and MSM patients at these health facilities (Figure 1). The study was approved by the Institutional Review Board of Albert Einstein College of Medicine (protocol number 2020-12619) and Ethical Committee of University of Kinshasa School of Public Health (protocol number ESP/CE/110/2021).

**Figure 1.**
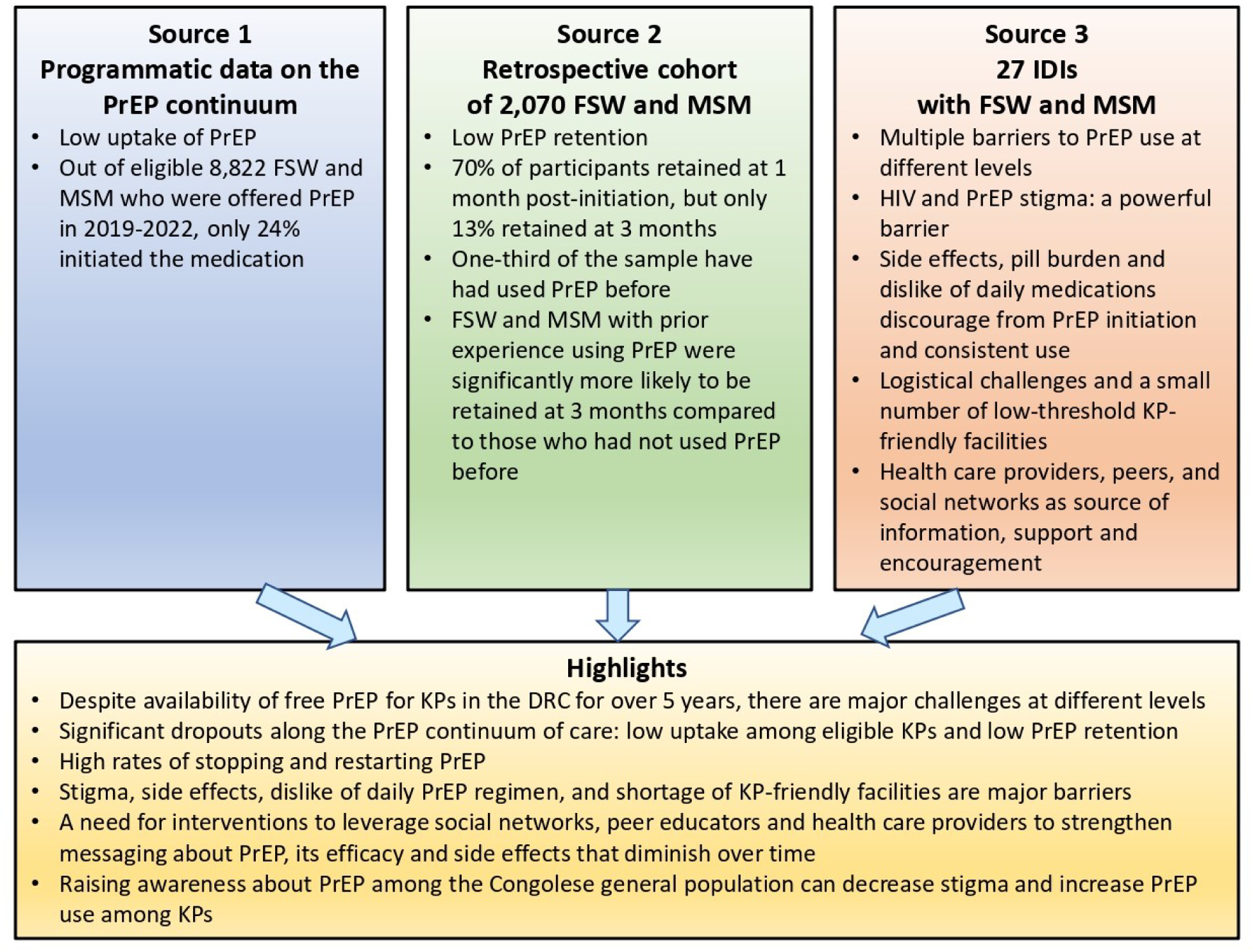
Joint display of the mixed-method study findings.

We conducted this study at five KP-friendly health facilities and drop-in centers (DIC) in Kinshasa, DRC, between 2019-2022. DRC is located in Central Africa and has a population of over 100 million. HIV prevalence among adult population is about 1% (22), but is two-or more times higher in the capital city of Kinshasa of 17 million people (23) and disproportionately higher among FSW (7.5%) and MSM (7.1%) (22). National guidelines recommend oral daily PrEP for individuals with elevated risk including sex workers, MSM, transgender people, injection drug users, and HIV-negative partners of people living with HIV. PrEP in the DRC is available free of change for these populations at risk. During an initial visit to a health facility (often for an STI assessment and/or treatment), health care providers share information about PrEP and screen patients for eligibility. KPs are also invited to come to DICs by peers. To be eligible for PrEP, patients must be aged 18 years or older, have recently tested negative for HIV, have normal estimated creatinine clearance, and be willing to commit to daily medication for HIV prevention. Individuals initiated on PrEP are seen for clinical visits and medication refills every three months.

### Data sources

We utilized data from three sources. Programmatic data on PrEP-related health service delivery was obtained directly from staff at the five participating sites. Second, the research team extracted routine clinical data from health center registries of FSW and MSM who initiated PrEP between 2019-2022 at these same five sites. Third, we conducted individual qualitative interviews with FSW and MSM patients at health centers.

### Data collection

To obtain programmatic data, we directly queried administrators or staff at each health center; they provided aggregate data on the number of individuals evaluated for, eligible, and initiating PrEP at each facility.

To form the retrospective cohort of FSW and MSM using PrEP, research staff extracted individual-level clinical data on patients initiating PrEP (including sociodemographic information, HIV risk, sexual health, and diagnostic testing) from clinical registries and inputted into an electronic Research data capture (REDCap) database.

Participants for semi-structured in-depth interviews (IDIs) were recruited using purposive sampling from the retrospective cohort of FSW and MSM. KP group (FSW and MSM) and PrEP status (currently using or stopped using) were used for purposive sampling. We also identified FSW and MSM who were evaluated for PrEP but never used it using clinical records at participating sites to enroll them in the study. From November 2021 through March 2022, trained research staff conducted 27 IDIs with FSW and MSM in French and Lingala to collect information about awareness of and willingness to use PrEP, experiences with PrEP, and barriers to and facilitators of PrEP use. A short questionnaire was administered at the end of IDIs to collect sociodemographic and health behavior information. IDIs were conducted in private rooms at participating clinics and DICs and were audio-recorded. Participants received 10,000 Congolese Francs (approximately 5USD) to compensate for their time.

### Quantitative variables and measurements

The primary outcome of interest was early PrEP retention, defined as attendance at scheduled appointments at one and three months after PrEP initiation. Sociodemographic data included age (18-30, 31-40, and >40 years) and marital status (single/divorced/separated vs married/cohabiting). Other covariates included estimated number of sexual partners in the past 6 months (0-89 vs 90+); regular condom use in the past 6 months (yes vs no); sex work being the principal source of income in the past 6 months (yes vs no); history of the previous PrEP use (yes vs no); and whether individuals had been diagnosed for an STI in the past 6 months (yes or no).

### Quantitative data analysis

First, we summarized programmatic data to create the PrEP cascade on numbers screened, eligible, and initiated on PrEP. Next, we analyzed the retrospective cohort data, stratifying all analyses by the KP groups – FSW and MSM – given their different characteristics, health behaviors, and potential factors associated with the use of PrEP and PrEP retention. Demographic and clinical characteristics of participants were summarized with proportions and counts. We calculated the proportion of participants who were retained at 1– and at 3-months among those who initiated PrEP. Logistic regressions were used to assess factors associated with early retention and to calculate odds ratios (ORs) and 95% confidence intervals (CIs) assessing the strength and direction of these associations.

### Qualitative data analyses

IDIs were audio-recorded and transcribed into text files in French or Lingala according to the interview language. Transcripts in Lingala were then translated into French for analyses. Using a combination of inductive and deductive approaches for thematic analysis, the research team conducted readings of five transcripts, meeting regularly to identify, discuss, and label common themes across transcripts. To broadly assess multi-level factors that could impact PrEP engagement, we developed the initial coding scheme using the socioecological model (24), then iteratively refined the codebook using consensus to resolve discrepancies. Four coders (AS, NM, NZ, and PL) then used DeDoose web-based software for qualitative analyses (25) to apply the codes to interview transcripts, with each transcript coded by at least 2 investigators. The team held regular meetings to review and discuss the coding process and resolve discrepancies if necessary. Upon completion of coding, we used the constant comparative method to identify, refine and consolidate emerging themes. We utilized Excel matrices to map themes by different levels of the socioecological model and to organize them by barriers and facilitators of PrEP use. Using the same principles of analyses, we also identified and examined differenced by KP group (FSW vs MSM) and by PrEP status (current users, stopped using, or never initiated PrEP).

### Data integration and synthesis

Integration in this MMR study occurred at different levels including the conceptual underpinnings of the study design and planning, data analyses, interpretation, and reporting of results (26). Integrated results were presented using a visual aid.

## RESULTS

### Quantitative results

From 2019 to 2022, the five study sites cumulatively screened 12,829 people for PrEP. 10,554 (82%) patients were HIV-negative: 4,788 (37%) FSW, 4,034 (31%) MSM, 1,703 (13%) clients of sex workers, and 11 (<0.1%) HIV-negative partners of PLWH. Out of 8,822 FSW and MSM screened eligible for PrEP, 2,070 (23.5%) initiated medication including 809 (39%) FSW and 1,261 (61%) MSM (Table 1). The majority of patients were aged 18-30 (77%, n=1,579) and single (99%; n=2,054). Most patients reported that they had not been using condom regularly (81%, n=1,670) and had high numbers of sexual partners (90+ partners in the past 6 months): 71% (n=568) of FSW and 36% (n=451) of MSM. Sex work was the principal source of income for 1,003 patients (49%). The large majority of patients (91%, n=1,871) had not been diagnosed with STIs in the preceding 6 months and 1,479 (71%) had not used PrEP before.

**Table 1.** Socio-demographics characteristics of FSWs and MSM who initiated PrEP in 2019-2021; n=2,070.

|  | **FSW, N (%)** | **MSM, N (%)** | **Total, N (%)** |
| --- | --- | --- | --- |
| Age |  |  |  |
| 18-30 | 561 (69.9) | 1018 (80.9) | 1,579 (76.6) |
| 31-40 | 194 (24.2) | 187 (14.9) | 381 (18.5) |
| >40 | 48 (6.0) | 53 (4.2) | 101 (4.9) |
| Marital status |  |  |  |
| Single/Divorced/Separated | 799 (99.3) | 1,255 (99.7) | 2,054 (99.5) |
| Married/cohabiting | 6 (0.7) | 4 (0.3) | 10 (0.5) |
| Number of sexual partners in the past 6 months |  |  |  |
| 0-89 | 232 (29.0) | 803 (64.0) | 1,035 (50.4) |
| 90+ | 568 (71.0) | 451 (36.0) | 1,019 (49.6) |
| Regular condom use |  |  |  |
| Yes | 283 (35.2) | 104 (8.3) | 387 (18.8) |
| No | 520 (64.8) | 1150 (91.7) | 1,670 (81.2) |
| Sex work being the principal source of income in the past 6 months |  |  |  |
| Yes | 732 (91.4) | 271 (21.6) | 1,003 (48.8) |
| No | 69 (8.6) | 985 (78.4) | 1,054 (51.2) |
| Used PrEP before |  |  |  |
| No | 528 (65.3) | 951 (75.4) | 1,479 (71.4) |
| Yes | 281 (34.7) | 310 (24.6) | 591 (28.6) |
| Had STIs in the past 6 months |  |  |  |
| Yes | 115 (14.3) | 66 (5.3) | 181 (8.8) |
| No | 692 (85.7) | 1179 (94.7) | 1,871 (91.2) |
| Retained at 1 month | 541 (66.9) | 929 (73.7) | 1,470 (70.8) |
| Retained at 3 months | 120 (15) | 121 (9.6) | 241(11.6) |

\* Frequencies may not add up due to missing data

Early retention on PrEP was low among both FSW and MSM, declining markedly by 3 months. Among 809 FSW, 79% (n=541) returned to the clinic for scheduled appointments and PrEP refills at 1 month after PrEP initiation, with only 15% (n=120) returning for a 3-month visit (Table 2). Similarly, among 1,261 MSM, 74% (n=929) were retained at 1 month following initiation of PrEP, with only 10% (n=121) returning for the 3 months follow-up (Table 3).

**Table 2.** Factors associated with PrEP retention at 1– and 3-month appointments among FSWs.

|  | Retained at 1 month (n=541, 66.9%) |  |  | Retained at 3 months (n=120, 15%) |  |  |
| --- | --- | --- | --- | --- | --- | --- |
|  | N | OR (95% CI) | aOR (95% CI) | N | OR (95% CI) | aOR (95% CI) |
| Age |  |  |  |  |  |  |
| 18-30 | 373 |  |  | 68 |  |  |
| 31-40 | 139 | 1.27 (0.89, 1.82) | 1.30 (0.90, 1.88) | 43 | 2.01 (1.29, 3.14) | **2.23 (1.30, 3.85)** |
| >40 | 26 | 0.60 (0.33, 1.08) | 0.66 (0.36, 1.22) | 9 | 2.37 (1.02, 5.55) | 2.51 (0.86, 7.27) |
| Number of sexual partners in the past 6 months |  |  |  |  |  |  |
| 0-89 | 163 |  |  | 12 |  |  |
| 90+ | 372 | 0.80 (0.58, 1.12) | 0.76 (0.52, 1.13) | 107 | 5.08 (2.71, 9.53) | **2.70 (1.24, 5.89)** |
| Regular condom use |  |  |  |  |  |  |
| Yes | 184 |  |  | 45 |  |  |
| No | 354 | 1.15 (0.84, 1.56) | 0.83 (0.56, 1.23) | 75 | 0.83 (0.54, 1.27) | 0.60 (0.33, 1.08) |
| Sex work principal source of income in the past 6 months |  |  |  |  |  |  |
| Yes | 493 |  |  | 94 |  |  |
| No | 46 | 0.97 (0.57, 1.64) | 0.96 (0.55, 1.66) | 25 | 5.05 (2.71, 9.41) | **3.73 (1.67, 8.34)** |
| Used PrEP before |  |  |  |  |  |  |
| No | 341 |  |  |  |  |  |
| Yes | 200 | 1.35 (0.99, 1.85) | 1.41 (0.99, 2.02) | 96 | 12.19 (7.40, 20.08) | **11.80 (6.39,21.79)** |
| Had STIs in the past 6 months |  |  |  |  |  |  |
| Yes | 62 |  |  | 10 |  |  |
| No | 478 | 1.91 (1.28, 2.85) | **1.78 (1.11, 2.85)** | 110 | 1.55 (0.76, 3.16) | 0.72 (0.29, 1.76) |

**Table 3.** Factors associated with PrEP retention at 1– and 3-month appointments among MSM.

|  | Return at 1 month (n=929, 73.7%) |  |  | Return at 3 months (n=121, 9.6%) |  |  |
| --- | --- | --- | --- | --- | --- | --- |
|  | N | OR (95% CI) | aOR (95% CI) | N | OR (95% CI) | aOR (95% CI) |
| Age |  |  |  |  |  |  |
| 18-30 | 745 |  |  | 91 |  |  |
| 31-40 | 144 | 1.23 (0.85, 1.77) | 1.27 (0.86, 1.87) | 24 | 1.44 (0.88, 2.35) | 1.14 (0.65, 2.00) |
| >40 | 38 | 0.93 (0.50, 1.71) | 0.97 (0.51, 1.87) | 6 | 1.35 (0.55, 3.31) | 0.78 (0.26, 2.41) |
| Number of sexual partners in the past 6 months |  |  |  |  |  |  |
| 0-89 | 614 |  |  | 68 |  |  |
| 90+ | 309 | 0.67 (0.52, 0.87) | 0.78 (0.59, 1.02) | 52 | 1.62 (1.10, 2.40) | 1.39 (0.90, 2.16) |
| Regular condom use |  |  |  |  |  |  |
| Yes | 71 |  |  | 17 |  |  |
| No | 852 | 1.33 (0.86, 2.05) | 1.36 (0.83, 2.22) | 103 | 0.44 (0.24, 0.78) | 1.40 (0.59, 3.32) |
| Sex work principal source of income in the past 6 months |  |  |  |  |  |  |
| Yes | 197 |  |  | 31 |  |  |
| No | 727 | 1.06 (0.78, 1.43) | 0.86 (0.62, 1.18) | 89 | 0.75 (0.48, 1.16) | 0.84 (0.52, 1.36) |
| Used PrEP before |  |  |  |  |  |  |
| No | 758 |  |  | 74 |  |  |
| Yes | 171 | 0.31 (0.24, 0.41) | **0.28 (0.21, 0.37)** | 47 | 3.50 (2.32, 5.29) | **4.08 (2.59, 6.41)** |
| Had STIs in the past 6 months |  |  |  |  |  |  |
| Yes | 36 |  |  | 12 |  |  |
| No | 877 | 2.42 (1.47, 4.00) | **3.02 (1.75, 5.23)** | 97 | 0.25 (0.12, 0.51) | **0.22 (0.10, 0.50)** |

Among FSW, those who had not been diagnosed with a sexually transmitted infection in the past 6 months, were significantly more likely to be retained at 1 month post-initiation compared to FSW who reported having STIs (aOR 1.78, 95% CI 1.11, 2.85). At 3 months, older age (31-40 vs 18-30; aOR 2.23, 95% CI 1.30, 3.85), a larger number of sexual partners (90+ vs 0-89; aOR 2.70, 95% CI 1.24, 5.89), and previous experience of using PrEP (yes vs no, aOR 11.80, 95% CI 6.39, 21.79) were significantly associated with retention on PrEP.

Among MSM, factors associated with retention at 1 month, were similar to those observed among FSW. Patients who had not been diagnosed with a sexually transmitted infection in the past 6 months, were significantly more likely to be retained at 1 month compared to MSM who reported having STIs (aOR 3.02, 95% CI 1.75, 5.23). At 3 months, previous experience using PrEP (aOR 4.08, 95% CI 2.59, 6.41) was significantly associated with retention.

### Qualitative results

From November 2021 to March 2022, we conducted 27 IDIs. Participants were between 22 to 52 years old and included 14 FSW (52%) and 13 MSM (48%). Among them, 14 individuals were using PrEP, 6 had previously been on PrEP but were not using at the time of the interview, and 7 had never used PrEP (Table 4). Most participants had completed secondary education (63%, n=17) and were single, divorced, or separated (85%, n=23).

**Table 4.** Characteristics of IDI participants (n=27)

|  | **FSW, N (%)** | **MSM, N (%)** |
| --- | --- | --- |
| Age |  |  |
| 18-30 | 6 (43) | 8 (62) |
| 31-40 | 3 (21) | 2 (15) |
| >40 | 5 (36) | 3 (23) |
| Marital status |  |  |
| Single/Divorced/Separated | 11 (79) | 12 (92) |
| Married/cohabiting | 3 (21) | 1 (8) |
| Education |  |  |
| Primary school | 2 (14) | 1 (8) |
| Secondary school | 10 (72) | 7 (54) |
| Some university | 2 (14) | 5 (38) |
| PrEP use |  |  |
| Current | 7 (50) | 7 (54) |
| Prior | 3 (21) | 3 (23) |
| Never | 4 (28) | 3 (23) |
| Sex at birth |  |  |
| Female | 12 (86) | 0 |
| Male | 2 (14) | 13 (100) |
| Gender identity |  |  |
| Female | 14 (100) | 0 |
| Male | 0 | 11 (85) |
| Gender fluid | 0 | 2 (15) |
| Sexual partners |  |  |
| Men | 14 (100) | 6 (46) |
| Men and women | 0 | 7 (54) |
| Number of sexual partners in the past 6 months |  |  |
| 1-20 | 3 (21) | 9 (82) |
| 20-99 | 1 (15) | 1 (9) |
| >100 | 9 (64) | 1 (9) |
| Had STIs in the past 6 months | 10 (71) | 8 (73) |
| Exchanged sex for money in the past 6 months | 14 (100) | 6 (46) |
| Reported challenges related to condom use |  |  |
| No challenges | 4 (33) | 2 (18) |
| Condoms feel uncomfortable and/or reduce pleasure | 2 (17) | 6 (55) |
| Condoms can tear | 4 (33) | 0 |
| Clients or partners unwilling to use condoms | 2 (17) | 3 (27) |
| Time since most recent HIV test |  |  |
| <1 month | 4 (33) | 2 (15) |
| 2-5 months | 7 (59) | 7 (54) |
| >6 months | 1 (8) | 4 (31) |

The sample of FSW included 12 individuals assigned female at birth and 2 transgender persons who identified as female. All reported exchanging sex for money in the prior 6 months and 10 (71%) reported having had an STI during this time. Two-thirds (n=9) of FSW reported having a large number of clients (400–600) over the prior 6 months.

Among MSM, 11 individuals identified as male and two as gender fluid. Almost half of MSM (46%, n=6) reported having exchanged sex for money in the 6 months prior to interviewing, and most (73%, n=8) reported having had an STI over this time period. The majority of MSM (82%, n=9) reported between 1 to 20 sexual partners in the 6 months prior to participation.

Our analysis revealed several emergent themes. Knowledge about PrEP protecting against HIV was high, and all participants knew that PrEP was available at KP-friendly health facilities and drop-in centers. Healthcare providers and social networks were the main sources of information, encouragement and support for PrEP use. These important facilitators notwithstanding, major barriers could outweigh benefits of using PrEP. Numerous concerns that discouraged FSW and MSM from PrEP initiation and consistent use included individual-level barriers such as side effects and pill burden, multiple stigmas operating at different levels, and logistical challenges such as distance to health centers.

### Side effects and pill burden: “Why take medications if I am not sick?”

Anticipated and experienced side effects proved to be a powerful barrier to PrEP use. Participants recounted a strong fear of side effects that discouraged some from PrEP initiation. Anticipated side effects were fueled by conversations with peers who heard about side effects of PrEP or experienced them first-hand. Misinformation, or exaggeration of possible side effects, spread within social networks of KPs, and served as an important barrier to medication use.

FSW and MSM who were currently using PrEP or discontinued medication, listed a range of experienced side effects. Fatigue and dizziness were most common, followed by nausea, headaches, heartburn, and excess salivation and urination.

> *“PrEP makes me really dizzy; it is like the earth is turning fast under my feet. I also had headaches at the beginning“*.
>
> *Female in their 30s, currently on PrEP*

Side effects became the primary reason for discontinuation of PrEP by both FSW and MSM. Pill burden, commonly expressed through the notion of “having had enough” or *“* having too much*“* PrEP, combined with side effects. These reasons were most often used to justify discontinuation of PrEP.

> *“I felt that it was becoming like a burden, taking it [PrEP] every day. I felt tired all the time, sometimes stomach aches, eh! I talked about it with him [doctor] at the beginning, I asked to stop, he reassured me by telling me, keep taking! But I couldn’t take it anymore, I gave up”*.
>
> *Female in their 40s, previously on PrEP*

> *“I stopped because I realized that I had consumed a lot. Sometimes, I had heartburns, eh! That’s why I stopped“*.
>
> *Female in their late 40s, previously on PrEP*

Large size of the pill was another concern, which further contributed to the dislike of PrEP:

> *“I stopped due to side effects that developed – nausea, vomiting – and the size of pills. The pills are big, really big. When you take it, you may believe that there is something stuck in your throat]. Eventually, my heart could not take it anymore*.
>
> *Male in their 20s, previously on PrEP*

Other factors such as daily routines and lifestyle could complicate consistent PrEP use. Several FSW indicated that experienced side effects, primarily fatigue and dizziness, made performing sex work more difficult. FSW prioritized sex work as their source of income, which made them seek solutions such as moving PrEP intake to the morning hours so that to have energy at night, or abandon the medication. Alcohol use was also an essential part of the FSW and MSM’s lifestyle who could drink with clients or socialize in bars and other venues frequented by KPs. Participants received mixed messages from healthcare providers who recommended not using alcohol while on PrEP or shared limited information on the combination of PrEP and alcohol. This discouraged some participants and complicated adherence to PrEP.

Side effects, pill burden, challenges to integrate PrEP into daily routines and other considerations intertwined with the general dislike of medications rooted in society and culture. Participants did not buy into the idea of daily pill intake and commonly demanded: “Why take medication if I’m not sick?” An idea of taking something on a daily basis seemed foreign for many participants, especially those who had never initiated PrEP. FSW and MSM alike did not necessarily specify their concern, providing instead vague explanations like “this is not for me”, “not now, but I may try one day” or similar. This FSW, who never initiated PrEP, shared a concise explanation of her unwillingness in strong terms of “addiction”:

> *“I don’t like something that demands so much from me. I mean, that I get addicted to it, take it every evening, because, they [doctors] say it’s to be taken every day and I don’t like taking [medical] products every day*”.
>
> *Female in their 20s, never used PrEP*

### HIV and PrEP stigmas

PrEP bottles look very similar to antiretroviral treatment (ART) taken for HIV. PrEP use thus informed many questions and rumors from family, friends and significant others, because PrEP was confused with ART. Daily use of medication was often believed to be associated with disease treatment, rather than taken for preventive purposes. Due to wide-spread stigma and confusion of PrEP with ART, participants preferred to take PrEP alone at home to maintain confidentiality and many FSW and MSM chose to not disclose PrEP use to anyone. However, disclosure may be unintentional in the context of small and often crowded living spaces. Family or friends could notice PrEP bottles, which caused a lot of doubt and questions. Anticipated stigma ruled out the possibility of PrEP initiation for some participants:

> *“One of the difficulties is that if you take PrEP in public, people will think you have HIV”*.
>
> *Male in their late 40s, never used PrEP*

> *“I refused to take PrEP because of the packaging; the bottle, essentially – the same bottle as ART“*.
>
> *Male in their 20s, never used PrEP*

In the context of powerful stigmas, operating at the individual, relational, and societal levels, hesitancy about the use of PrEP and decisions about its (dis)continuation were often rooted in the perceived balance of benefits and negative outcomes, and the dynamics of relationship with others. Participants who emphasized that they put their health and protection against HIV first, like this participant, adhered to PrEP despite stigma:

> *“People were saying: “He takes medicine for people with AIDS !“ But I didn’t care much“. Male in their 20s, currently on PrEP*

Other participants had a difficult time and, despite appreciation of PrEP as efficient protection, eventually discontinued due to side effects and stigma, enacted at different levels:

> *“First of all, I was ashamed because I heard from others all the time that this same medicine that we take, it seems that it is the same that the sick [with HIV] take. I was ashamed even at home, my family suspected me. They thought I wasn’t clean; that when you start taking big tablets like that, you have [HIV], or something like that, but I held on. Even when I came to the center to take my medicine, you see how they look at me. That’s what bothers me, but it was my goal and I continued, but eventually stopped“*.
>
> *Male in their 20s, previously on PrEP*

Most participants acknowledged lack of information or misinformation about PrEP among KPs and broader Congolese society, which contributed to PrEP use stigma. Participants noted a lack of awareness about PrEP that caused doubts about its effectiveness and reinforced negative labeling of PrEP users and perpetuated HIV stigma. They suggested that raising awareness about PrEP could improve its acceptance in the society and potentially decrease stigma.

### Accessibility of PrEP

Finally, logistical barriers and PrEP being available in a limited number of KP-friendly health centers and DIC were also important barriers to PrEP uptake and adherence. Participants brought up the problem of a long distance between facilities offering PrEP services and their homes. With an estimated population of 17 million, Kinshasa is the most populous city in the DRC with significant traffic and lack of public transportation, making travelling between different city’s areas complicated and time-consuming.

> *“A difficulty that people may encounter is the distance; the journey to the center is quite long“*.
>
> *Female in their 20s, never used PrEP*

Combined with possible PrEP stockouts at health facilities and DIC, the problem of PrEP accessibility is stark. Participants demonstrated a strong preference for developing more centers offering PrEP services across the city.

> *“I think that we need to set up several centers in the city. That will make it easier for everyone to access from their homes“*.
>
> *Male in their late 40s, never used PrEP*

### “It’s a good product!” Experiences using PrEP and facilitators at different levels

Perceived balance of positive and negative outcomes associated with PrEP use differed among FSW and MSM who discontinued or never initiated PrEP, and participants who had been using it at the time of research. Pervasive barriers, primarily side effects and stigma, hindered initiation of PrEP or forced participants to stop taking the medication. Although all participants had a good understanding of risks stemming from their sex work or behaviors, current PrEP users brough protection against HIV to the forefront. They emphasized efficacy of PrEP in protecting them against HIV, which made them feel at ease in the context of multiple partners, violence against FSW, and condomless sex.

> *“From my experience, I can say that PrEP is effective, because I don’t use condoms, I don’t like condoms. Since I started taking PrEP, I don’t really fear about getting HIV”. Male in their 20s, currently on PrEP*

Among participants, who persisted on PrEP, initial side effects gradually weaned off as participants got used to PrEP and integrated it into daily routine. Other important facilitators also existed on the relational, social, and institutional levels. Social networks were essential in providing information and support for both FSW and MSM. Networks included members of close-knit communities of FSW, MSM and transgender persons, peer educators at drop-in centers and health facilities, and KP community centers. These were powerful sources of information, advice and encouragement for PrEP uptake and use. Participants who learned first-hand about PrEP efficacy and felt comfortable using, talked to peers and encouraged them to try PrEP.

> *“I have already shared my story with many people and told them to use PrEP! I told my friends here and in other neighborhoods; I invited them to come [to the health center]. Because without PrEP it’s difficult, really, especially with our lifestyle. PrEP protects us a lot; without that I could have HIV”*.
>
> *Transgender female in their 20s, currently on PrEP*

Spreading information about PrEP, participants raised awareness and encouraged friends to try this “good product”. Although in a large city of Kinshasa at the intersection of gendered sex work and migration many KPs face logistical barriers to access PrEP, interest in PrEP was high. Opening up to peers and significant others (*cheris,* permanent partners) allowed participants to discuss PrEP and benefit from increased support and encouragement.

Healthcare providers, peer educators, and organization of health systems served as essential institutional facilitators of PrEP use. FSW and MSM valued KP-friendly health centers and drop-in community centers (DIC) as the best place to receive information about PrEP and access services. Participants enjoyed welcoming positive attitudes of healthcare providers who educated them about PrEP and its availability, and offered a full range of STIs and HIV testing and PrEP services. Participants also appreciated confidentiality provided by health facilities centers and DICs. Although DICs were located in residential neighborhoods, frequented by KPs, they usually had no signs indicating what population groups they served or which services provided. This provided participants with the needed anonymity and ensured trust in healthcare providers and peer educations who worked at these facilities. A good organization of the continuum of sexually transmitted infections (STIs) STI, HIV, and PrEP services contributed into participants displaying a strong preference for the continued provision of PrEP at the same facilities rather than pharmacies and other options discussed during interviews.

## DISCUSSION

Integrated findings from this mixed methods study highlight specific barriers among FSW and MSM in the DRC that ultimately led to low uptake and retention. Only one-quarter of FSW and MSM who were screened and deemed eligible for PrEP ultimately initiated it (Figure 1; Source 1). Similarly, while approximately 70% of FSW and MSM who were initiated on PrEP returned for scheduled clinical visits and PrEP refills at 1 month, only 15% of FSW and 10% of MSM were retained at 3 months. These findings are consistent with evidence from other African settings, which showed that retention on PrEP falls significantly by 12 months (14-18; 27,28). However, continuation rate at 1– and 3-months in our Congolese FSW and MSM samples was very low compared to other studies. A systematic review of PrEP uptake, retention and adherence among FSW in SSA found that pooled retention on PrEP at 6 months was 66% and 83% for facility-based and community-based delivery models, respectively (16). There is mixed evidence on PrEP retention among MSM in SSA, pointing out that retention at 6 months ranged from 30% in Kenya (14) to 88% in Benin (29).

FSW and MSM who had prior experience using PrEP were significantly more likely to be retained by 3 months compared to those who had not used PrEP before. Almost one-third of participants in the retrospective cohort (Figure 1; Source 2) had previously used PrEP, while percentage of prior users was higher among FSW (35%; n=281) compared with MSM (25%, n=310). Relatively high rates of prior PrEP use are consistent with evidence from SSA and global settings. A systematic review found that, overall, 24% of people who had stopped PrEP, restarted using the medication. Restarting of PrEP was higher in studies from Africa compared with evidence from the USA (30). A prospective cohort study of MSM and transgender women from Kenya found that over the course of the 20-month follow-up, 40% of participants stopped PrEP and, of these, 51% restarted PrEP (27). These findings from diverse settings suggest that people might need a few attempts before successfully fully engaging on PrEP. People may also have different use patterns associated with changing risk of HIV exposure and thus need flexibility in being able to easily stop/restart as clinically appropriate. Importantly, removal of barriers to access the medication was found to be a reason for restarting PrEP (30).

Qualitative findings from this study highlighted important barriers that may explain observed patterns of low PrEP initiation and retention, and shed light on restarting PrEP. Experienced and anticipated side effects, pill burden, and challenges associated with daily pill intake were essential individual-level barriers. Stigma associated with PrEP and PrEP users due to similarity of PrEP to ARVs were another essential barrier to PrEP adoption and use among FSW and MSM. Participants faced suspicion and judgment from those around them, which led to difficulty maintaining confidentiality while using PrEP. The confusion of PrEP with ARVs contributed to stigma by creating suspicion and unjustified fears of participants’ being HIV-infected among family, friends, and peers. Even individuals who acknowledged PrEP efficiency and committed to its continued use often found it challenging due to combined effects of stigma and peer– and family pressure of suspicion and doubts. These findings are consistent with global evidence that identifies PrEP stigma as a major factor that significantly impedes PrEP acceptability, uptake, and adherence by affecting acceptance at individual-, provider-, and community-levels (31–37).

Culturally rooted dislike of daily medication use outside of the illness context is another factor affecting PrEP initiation and use. Globally, research has long shown the role of cultural factors behind concerns over daily medication intake that affect adherence including antiretroviral treatment for HIV (38). Although there is lack of data from African settings on cultural attitudes towards daily oral PrEP, our findings suggest the need to consider prejudice against daily medication use as an essential barrier.

Addressing these barriers may help to increase PrEP use among KPs at high risk of HIV acquisition in the DRC. Our study identified important facilitators of PrEP use that can potentially offset stigma and individual-level barriers. Health care providers, peer educators and social networks of FSW and MSM were essential in raising awareness and spreading information about PrEP. This was consistent with evidence from the DRC and Rwanda on the important role of health care providers who provide information about PrEP and build rapport with KPs during clinical visits and, as a result, facilitate initiation of PrEP (17,19). Similar to an early study from the DRC (19), we found that positive non-stigmatizing attitudes of providers are key to individuals’ decisions to initiate PrEP. Studies from Nigeria and Zimbabwe have shown that KPs endorsed introduction of PrEP within familiar settings at health facilities and DICs and appreciated encouragement from clinical staff and peers, which could overweigh stigma and the fear of side effects (39,40). Our findings suggest that PrEP can be provided for MSM and FSW through the same facilities in Kinshasa, DRC, but their number should be significantly increased to expand access to individuals at high risk of HIV across the large city. Programs to strengthen messaging from health care providers and peer educators could encourage sustained engagement in PrEP, particularly in the context of high rates of stopping and restarting PrEP. Programs to raise awareness among the general population are also needed to prevent stigmatizing labeling of PrEP users as HIV-infected. Finally, long-acting injectable PrEP has the potential to counter overlapping HIV and PrEP stigma and difficulty taking daily medication. Although long-acting PrEP has not yet been registered and introduced in the DRC, PEPFAR plans on the fast roll-out of injectable PrEP in other SSA countries. With a potential high demand among Congolese FSW and MSM, there is a need for policies to introduce long-acting PrEP in the DRC.

Several limitations of this study should be acknowledged. The study occurred in Kinshasa, the capital of the DRC. Participants recruited in Kinshasa may not represent experiences of key population in rural areas of the DRC. Willingness to participate in IDIs could be impacted by social desirability bias: some participants could be too stigmatized to participate in research about PrEP, or were unwilling to share their experiences during interviews conducted at health care facilities of DICs. We therefore may not have captured perspectives of KPs who had negative experiences interacting with health care settings to access PrEP services.

## CONCLUSIONS

Findings from this mixed methods study showed low rates of PrEP initiation and retention among Congolese FSW and MSM. Despite availability of PrEP in the DRC for over 5 years, major barriers must be addressed to increase PrEP use among KPs and combat HIV acquisition.

Primary barriers included side effects, the conflation of the HIV and PrEP stigma, and negative attitudes to daily PrEP regiment. This suggests the need to leverage social networks, peer educators and health care providers to strengthen messaging about PrEP, its efficacy and side effects that diminish over time. Raising awareness about PrEP among the Congolese general population can help to avoid negative labeling of PrEP users and improve PrEP acceptance and retention among key populations at risk.

## Data Availability

The datasets analyzed during the current study are available from the corresponding author on reasonable request.

## DECLARATIONS

### Ethical approval and informed consent

The study was approved by the Institutional Review Board of Albert Einstein College of Medicine (protocol number 2020-12619) and Ethical Committee of University of Kinshasa School of Public Health (protocol number ESP/CE/110/2021). All research participants provided written informed consent prior to in-depth interviews.

### Consent for publication

Not applicable

### Authors contributions

NZ and AS oversaw data collection and led data analyses, with support from QS, JR, and VP. JR, AA and VP conceived of and led the study. AS, PL, and NM were responsible for data collection and contributed to data analyses. KA, MY and AA contributed to interpretation of study data. DK and PN contributed to study design. All authors read and approved the final manuscript.

## Acknowledgments

We thank all those who participated in this study. We are grateful for the dedication and effort of the Congolese team that made this research possible. We thank the staff of the participating clinics and health authorities in the DRC.

